# Cerebrospinal fluid and brain α-synuclein seed amplification in autopsy-confirmed Lewy body disease relates to the distribution of pathology

**DOI:** 10.1101/2022.02.28.22271232

**Authors:** Moriah R. Arnold, David G. Coughlin, Barbara H. Brumbach, Denis S. Smirnov, Luis Concha-Marambio, Carly M. Farris, Yihua Ma, Amprion Inc., Yongya Kim, Jeffrey A. Kaye, Annie Hiniker, Randy L. Woltjer, Doug R. Galasko, Joseph F. Quinn

**Affiliations:** Medical Scientist Training Program, Oregon Health and Science University; Department of Neurosciences, University of California San Diego; Biostatistics and Design Program, Oregon Health and Science University; Amprion Inc; Department of Neurology, Oregon Health and Science University; Department of Pathology, University of California San Diego; Department of Pathology, Oregon Health and Science University; Portland VA Medical Center, Parkinson’s Disease Research Education and Clinical Care Center (PADRECC)

## Abstract

**Objective:** To determine the sensitivity and specificity of α-synuclein seed amplification assay (αSyn-SAA) in antemortem and postmortem CSF and brain homogenate samples of autopsy-confirmed patients with a spectrum of Lewy-related pathology (LRP).

**Methods:** Antemortem CSF samples were examined from 119 subjects with standardized neuropathological examinations from OHSU and UCSD (56 additional postmortem CSF samples available). The assay was also applied to frontal cortex and amygdala tissue to determine if the results could be explained by a regional variation in the propensity for seed aggregation. Sensitivity, specificity, and assay kinetics were compared across pathology groups and clinical data was compared across αSyn-SAA positive and negative groups.

**Results:** Fifty-three LRP-individuals and 66 LRP+ individuals (neocortical (n=38), limbic (n=7), and amygdala-predominant (n=21)) were included. There was a sensitivity of 97.8% and specificity of 98.1% of the αSyn-SAA to identify patients with limbic/neocortical pathology from antemortem CSF. Sensitivity to detect amygdala-predominant pathology was only 14.3%. Postmortem CSF and brain tissue αSyn-SAA analyses showed a similar detection pattern, with higher positivity in samples from limbic/neocortical cases. Kinetic parameters of aggregation were significantly slower in amygdala-predominant cases compared to limbic and neocortical cases.

**Interpretation:** In this multicenter study of autopsy-confirmed subjects with a spectrum of Lewy-related pathology, we confirm that the αSyn-SAA using CSF and brain tissue reliably identifies α-synuclein seeds in patients with diffuse pathology and related cognitive symptoms. Pathological α-synuclein in the amygdala appears less likely to form detectable seeds, which may result from differences in abundance, conformation, or strains of α-synuclein.

**Summary for Social Media If Published:**

1. Twitter handles of the authors: none
2. Alpha-synuclein seed amplification assays have shown high sensitivity and specificity in clinically defined DLB and PD cohorts
3. It is less well known how well these assays detect synuclein seeds across a pathologically defined spectrum of Lewy body disease. Here we examine the ability of the αSyn-SAA to detect alpha-synuclein seeds in a multicenter cohort of autopsy-validated cases with a spectrum of Lewy body related pathology.
4. High sensitivity and specificity of the αSyn-SAA is confirmed in detecting alpha-synuclein seeds in spinal fluid and brain tissue in limbic and neocortical stage Lewy body stage pathology, but markedly decreased sensitivity is observed in detecting alpha-synuclein seeds in both spinal fluid and brain tissue in amygdala-predominant type Lewy body related pathology. A small number of these cases showed seeding capability from the amygdala that was not present in the frontal cortex, suggesting a topographic spread of alpha-synuclein seeds.
5. The current generation of αSyn-SAAs have a high sensitivity and specificity for detecting the most clinically relevant forms of Lewy body related pathology. Further study is needed to understand the differences in Lewy body related pathology between limbic/neocortical cases and amygdala-predominant cases that result in this difference in seeding capability.

## Introduction

Aggregated α-synuclein (αSyn) is the main component of cytoplasmic inclusions called Lewy bodies (LB) and Lewy neurites (LNs), which are the defining pathological features of Parkinson’s disease (PD) and dementia with Lewy bodies (DLB)^1, 2^. In addition, αSyn-laden LBs are found in the brains of as many as 50-60% of sporadic Alzheimer’s disease (AD) cases^3-7^, 96% in familial *PSEN1* cases^8^, and in 10-20% of normal elders^9, 10^. The distribution of Lewy-related pathology (LRP) in AD tends to affect the amygdala and limbic regions as opposed to the caudo-rostral spread observed originally by Braak et al. in PD and DLB^8, 11^. Studies suggest that Alzheimer’s disease with amygdala-predominant Lewy bodies (AD/ALB) may have an earlier onset and steeper rate of decline in various clinical measurements when compared to AD without ALB, but are unlikely to result in a DLB clinical phenotype^12, 13^. Neuropathological analysis of DLB cases revealed numerous unique αSyn pathologies within the amygdala, such as neuropil αSyn aggregates, astrocytic αSyn, and carboxy-truncated forms of αSyn, which are highly prone to aggregation; however, many of these species were observed to a much lesser extent in AD/ALB cases^14^.

To date, neuropathological assessment at autopsy remains the gold standard to diagnose these diseases and *in vivo* αSyn biomarkers have been an unmet need. Recently, αSyn Seed Amplification Assays (SAAs), such as protein misfolding cyclic amplification (PMCA) and real time quaking induced conversion (RT-QuIC), have been adapted to detect misfolded αSyn aggregates (αSyn seeds) in CSF and peripheral tissues with remarkable diagnostic accuracy^15-20^. αSyn-SAA in CSF of clinically diagnosed PD and DLB patients has shown impressive results, with several independent groups reporting sensitivities and specificities near or above 90%^15-17, 21, 22^. However, detailed studies of αSyn-SAA performance in neuropathologically validated cohorts with varying distribution of LRP and clinical diagnoses other than LB disease are rare^23^.

Thus, it remains to be seen if differences in seeding activity are associated with different types or distributions of LRP. While it has recently been demonstrated that αSyn seeds associated with MSA likely have different αSyn seeding properties than PD/DLB^24^, less is known about the potential differences in seeding activity in the setting of AD/ALB. A few small studies have reported detecting aggregated αSyn seeds in CSF from clinically diagnosed AD patients (5/14 or 36% in one report^16^ and 0/16 in another^17^) and from patients clinically diagnosed with AD who were pathologically confirmed to have DLB (11/17 or 65%) or incidental Lewy bodies (2/13 or 15%)^15^. Together, this suggests that current assays may have different sensitivities for certain LRP αSyn species or that the propensity of pathological αSyn to seed may vary in PD and DLB compared to AD/ALB.

In this multi-center study, we evaluated the capability of αSyn-SAA to detect αSyn seeds in antemortem and postmortem CSF samples as well as brain tissue of patients who underwent autopsy and neuropathological analyses. We compared the αSyn-SAA results to clinical and neuropathological data to determine sensitivity, specificity, clinical, and pathological correlations of this assay across a spectrum of LRP.

## Methods

### Patient samples

Participants in brain aging studies from the Oregon Alzheimer’s Disease Center (OADC) and University of California San Diego Shiley-Marcos Alzheimer’s Disease Research Center (UCSD-ADRC) who had 1) CSF collection during life, and 2) subsequent brain autopsy (n=119) were included in the study. All subjects had an annual battery of clinical, neuropsychologic, and other cognitive assessments, as described by the National Alzheimer’s Coordinating Center (NACC)^25^, including Mini-Mental State exam (MMSE), and Unified Parkinson’s Disease Rating Scale Part III (UPDRS). Blood was drawn for the determination of APOE genotype. Clinical diagnoses, assigned at the time of CSF collection, included AD (n=75), DLB (n=9), PD (n=4), mild cognitive impairment (MCI, n=11), other dementia (n=13, including frontotemporal dementia (n=10), mixed dementia (n=1), and “other dementia” (n=2)), and cognitively normal controls (n=7). Clinical diagnoses were assigned by a multidisciplinary consensus conference at each site. CSF was collected by lumbar puncture in the morning fasting condition according to a standardized protocol^26^. A subset of patients (n=56) had additional CSF samples obtained at the time of brain removal. CSF specimens were divided into 0.5 ml aliquots and stored at −80°C until thawed for αSyn-SAA analysis. Antemortem CSF collection occurred 1-15 years prior to autopsy (average of 4.9 years).

Neuropathological assessments were performed in a standardized manner with various pathologies assessed using hematoxylin and eosin staining and immunohistochemistry directed against tau, amyloid-β, α-synuclein, and TDP-43 species as appropriate and pathological diagnoses were assigned by expert neuropathologists^27-31^. MSA cases were excluded from this study given the known altered kinetics on αSyn-SAA assays compared to PD and DLB cases^24^. Alzheimer’s disease neuropathological change was assigned according to NACC guidelines after Braak tau stage, CERAD stage, and Thal phase was determined^28, 32, 33^. Distribution of LRP was determined via α-synuclein immunohistochemistry staining (OADC: αSyn MJFR1, Abcam; UCSD-ADRC: pSer129 αSyn 81A, Biolegend Laboratories) using slices from pons and/or midbrain, hippocampus, amygdala, and neocortical areas including temporal cortex and/or middle frontal cortex and the following staging definitions were applied: Neocortical: midbrain+ pons+ hippocampus+ amygdala+ neocortex+; Limbic: midbrain+ pons+ hippocampus+ amygdala+ neocortex-; Amygdala-predominant: midbrain-pons-hippocampus+/- amygdala+ neocortex-^34^.

In a subset of patients (n=22), 500mg samples of frozen brain tissue from the middle frontal cortex and amygdala were provided for αSyn-SAA. Cases were selected that represented a spectrum of LRP: LRP-(n=4), amygdala-predominant LRP (n=10), and limbic/neocortical LRP (n=8). All frozen samples were provided from the UCSD-ADRC where right hemibrains had been sectioned and frozen at −80°C immediately post-autopsy for future biochemical analyses.

### Brain tissue homogenization

Frontal cortex and amygdala samples were homogenized to 10% w/v in 1XPBS (Cytiva, cat# SH30256.02) with cOmplete Mini EDTA-free protease inhibitor cocktail (Roche, cat# 11836170001). Approximately 100µg of brain sample was homogenized in 1.5mL tubes preloaded with 1mm zirconium beads (cat# 11079110zx) in a MP FastPrep 24 homogenizer. Two rounds of homogenization were performed for all samples (15s at 4m/s and 30s at 6m/s). If additional homogenization was needed, samples were chilled on ice for 5min in between additional homogenization rounds at 6m/s for 30s. BHs were centrifuged at 800xg for 1 minute to remove cellular debris. Supernatants were collected, vortexed, aliquoted, and stored at −80°C until αSyn-SAA analysis. BH aliquots were 10-fold serially diluted in synthetic CSF (Amprion, cat# S2022) up to 10^−8^ and analyzed in triplicates.

### αSyn Seed Amplification assay

CSF samples were initially analyzed by the endpoint qualitative version of the αSyn-SAA that has been validated for clinical use under CLIA/CAP certifications (clinical assay, SYNTap™). Each sample was analyzed in triplicate (40µL CSF per well) in a 96-well plate (COSTAR, cat# 3603). The reaction mixture consisted of 0.3mg/mL rec-αSyn (Amprion, ca# S2021) in 100mM PIPES pH 6.50, 500mM NaCl, 10µM ThT, and a 2.5mm borosilicate glass bead per well. This clinical version of the assay was performed according to standard operational procedures in agreement with CLIA regulation. CSF samples were deemed “Detected” or “Not Detected” based on a preestablished threshold for the median maximum fluorescence of the triplicate. The research and development (R&D) kinetic αS-SAA was utilized to analyze CSF samples and brain tissues. The methods of the kinetic αS-SAA have been reported in detail elsewhere^21, 22^. Briefly, CSF samples and brain homogenates (BHs) were evaluated in triplicates (40µL/well) in a 96-well plate (COSTAR 96, cat# 3916), in a reaction mix consisting of 0.3mg/ml rec-αSyn (Amprion, cat# S2021), 100mM PIPES pH 6.50 (Sigma, cat# 80635), 500mM NaCl (Lonza, cat# 51202), 10µM ThT (Sigma, cat# T3516), and a 3/32-inch BSA-blocked Si_3_N_4_ bead (Tsubaki Nakashima). This assay was performed in a BMG FLUOstar Omega shaker/reader that allows high frequency agitation at 37°C of a single plate with automatic fluorescence readings every 30min for accurate estimation of kinetic parameters. The assay outcomes of the R&D kinetic assay are positive, inconclusive, or negative, based on a probabilistic algorithm that uses maximum fluorescence and kinetic parameters^21^. Maximum fluorescence (F_max_, RFU) was the highest fluorescence reading within the length of the assay. A 4-parameter fit (Mars, BMG) was fit to estimate the slope (RFU/h) and the time to reach 50% of the F_max_ (T_50_, h) of each replicate/well. The time to threshold (TTT, h) was determined with a user defined formula (Mars, BMG); threshold was set to 5,000 RFU. Scientists performing the assay were blinded to the clinical or pathological diagnoses associated with the samples.

### Statistical Analysis

Clinical and pathological differences between the OADC and UCSD-ADRC cohorts were assessed to determine the necessity for stratification by site. All DLB and PD patients were from UCSD. Sensitivity, specificity, and predictive values were calculated via Chi-square test with 95% confidence intervals calculated using the hybrid Wilson-Brown method. Differences in kinetic parameters were analyzed by one-way ANOVA with Tukey’s multiple comparisons test or unpaired t test. Prior to testing group differences, all outcome variables were assessed for normality. For normally distributed continuous variables, we used the General Linear Model (GLM) to test whether there were group differences in the outcome variables (age at death, onset of cognitive symptoms, and MMSE decline rate). For non-normally distributed continuous variables (UPDRS at lumbar puncture, MMSE at lumbar puncture, UPDRS at most recent visit, MMSE at most recent visit, CDR at most recent visit, lumbar puncture to autopsy interval, amyloid beta protein in antemortem CSF, tau in antemortem CSF), we used a Kruskal-Wallis test (more than two groups) or a Wilcoxon rank-sum test (two groups) to test for group differences. Post hoc pairwise comparisons were tested using the Dwass, Steel, Critchlow-Fligner Method. We used chi-square tests to test for group differences when outcome variables were categorical (biological sex, early-onset status, clinical diagnosis, NACC variables: Thal phase for amyloid plaques (NPTHAL), Braak stage for neurofibrillary degeneration (BRAAK), density of neocortical neuritic plaques (CERAD), NIA-AA Alzheimer’s disease neuropathologic change (ADNC), density of diffuse plaques (CERAD-semi), cerebral amyloid angiopathy (NACCAMY), arteriosclerosis (NACCARTE), APOE status). For the following variables, we had data from both the OADC and UCSD-ADRC cohorts: age at death, rate of MMSE decline, MMSE at lumbar puncture, most recent MMSE score, interval between lumbar puncture and autopsy, biological sex, clinical diagnosis, THAL, BRAAK, CERAD, ADNC, APOE genotype. The following variables were only available from the OADC cohort: onset of cognitive symptoms. UPDRS score at lumbar puncture was only collected at UCSD. Statistical significance was set at *p* < 0.05.

## Results

### Neuropathological αSyn analysis and comparison

The neuropathological analysis of the 119 subjects revealed LRP in the brains of 66 (55%). Of the 66 LRP+ patients, 38 showed neocortical stage LRP, 7 showed limbic stage LRP, and 21 showed amygdala-predominant LRP. Given the low number of limbic stage LRP cases, limbic and neocortical cases were analyzed as a combined group for subsequent analyses.

Using a Kruskal-Wallis test, we compared patients within synuclein distribution groups (LRP-, amygdala-predominant, limbic/neocortical) on several standardized clinical and pathological variables to determine if there were important group differences. UPDRS part III scores were significantly different between LRP groups at lumbar puncture (*X*^*2*^=21.59, p<0.0001, Table 1) and at last visit prior to death (*X*^*2*^=14.93, p=0.0006, Table 1). Post-hoc analyses showed that the limbic/neocortical group had higher UPDRS part III scores at lumbar puncture than the LRP- and amygdala-predominant groups (Wilcoxon z=-3.71, p=0.0006 and Wilcoxon z=-3.44, p=0.002 respectively, Fig 1A). The limbic/neocortical group also had higher UPDRS III scores at last visit prior to death compared to the amygdala-predominant group (Wilcoxon z=-3.70, p=0.0007, Fig 1B). The majority of patients diagnosed with DLB (8/9) and PD (4/4) showed limbic/neocortical LRP rather than amygdala-predominant LRP, while 16/21 amygdala-predominant patients had a clinical diagnosis of AD (*X*^*2*^=28, p=0.002, Table 1, Fig 1C). Lastly, male sex was over-represented across the three synuclein distribution groups (*X*^*2*^=6.94, p=0.03, Table1).

**Table 1.**
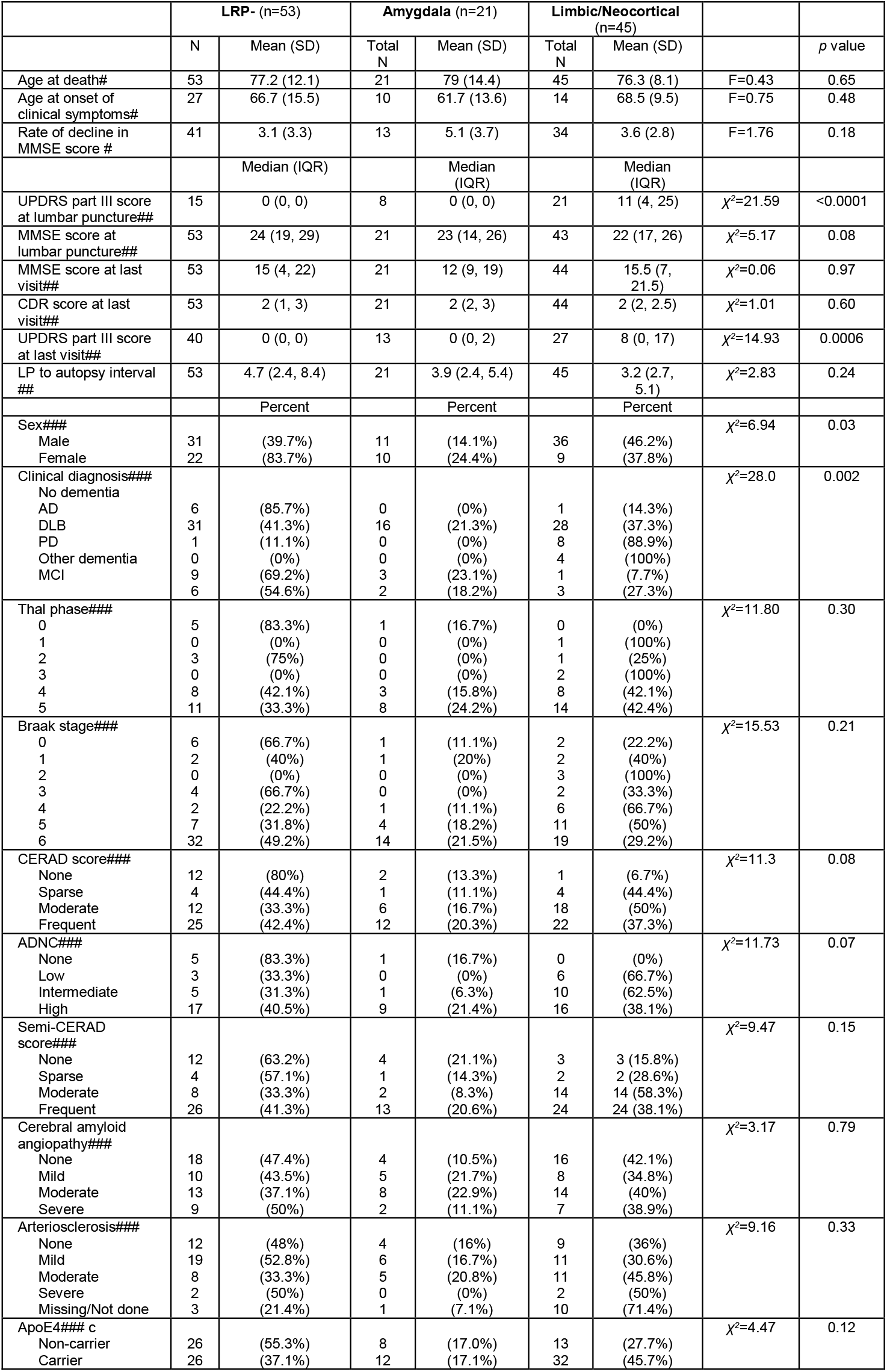
Summary of statistical analysis. # general linear model used for analysis, ## Kruskal-Wallis test used for analysis, ### Chi-square test used for analysis.

**Figure 1.**
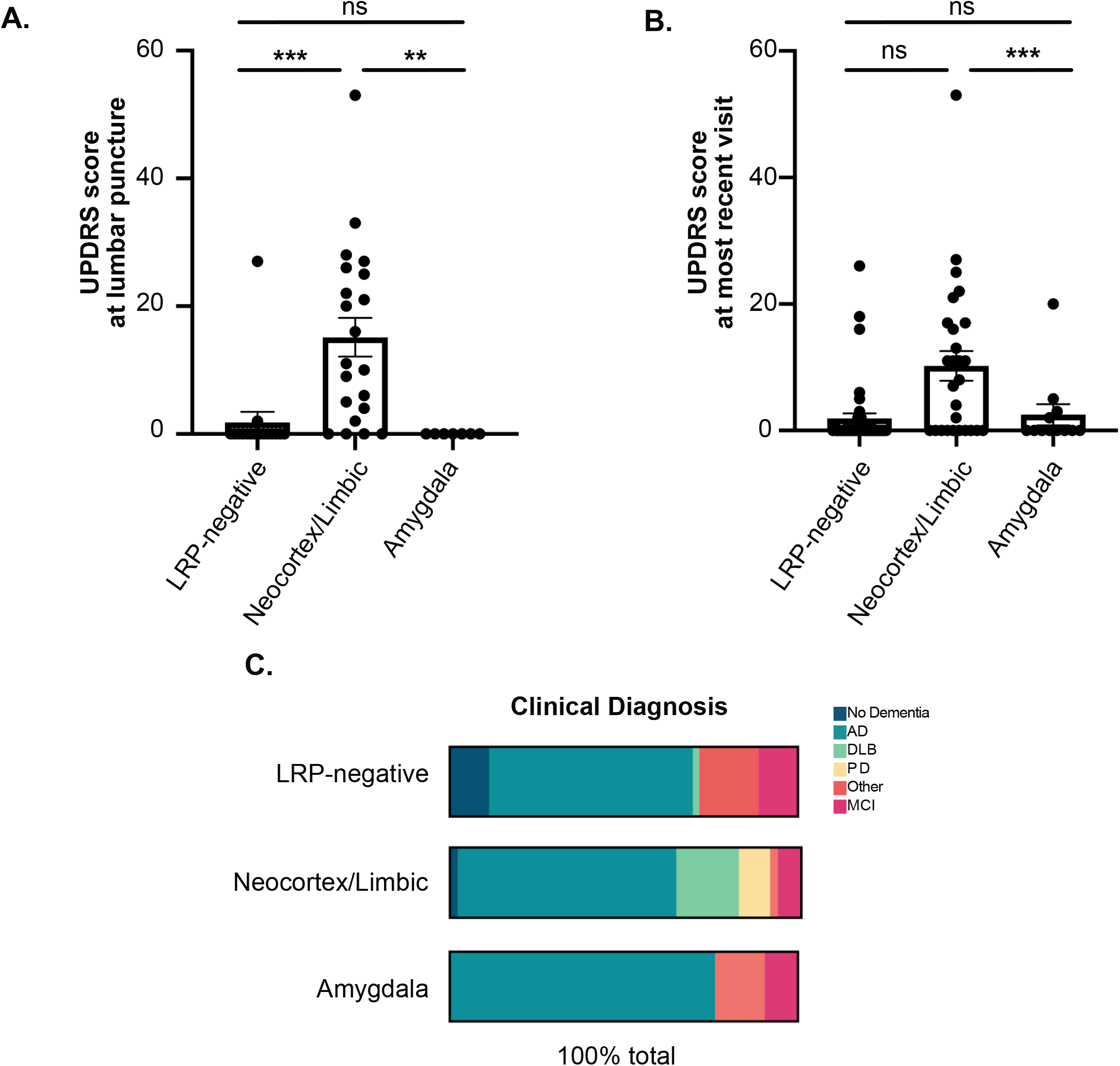
Clinical and pathological differences between LRP+ and LRP- groups stratified by alpha-synuclein distribution. A) UPDRS score at lumbar puncture and B) at most recent visit prior to death between LRP-negative, Neocortex/Limbic, and Amygdala-predominant groups. C) Distribution of clinical diagnosis in LRP-negative, Neocortex/Limbic, and Amygdala-predominant groups (AD=Alzheimer’s Disease, DLB=Dementia with Lewy bodies, PD=Parkinson’s Disease, Other=Other forms of dementia, MCI=Mild Cognitive Impairment). Statistical analysis using Wilcoxon rank-sum test with post hoc pairwise comparisons from Dwass, Steel, Critchlow-Fligner method (A, B) or chi-square (C). Error bars represent Standard Error of the Mean (SEM).

### Sensitivity and specificity of the αSyn-SAA using CSF samples

A total of 119 antemortem CSF samples were analyzed with αSyn-SAA. All but 1 of the 53 LRP-patients were negative by the clinical αSyn-SAA and thus, the specificity for the clinical assay in this cohort was 98.1% (95% CI 90.1% to 99.9%) (Table 2). Of the 66 LRP+ individuals, 47 were found positive by the clinical αSyn-SAA. The overall sensitivity of the assay to detect LRP in any form was 71.2% (95% CI 59.4% to 80.7%). However, significant differences were observed when stratifying sensitivity analysis by LRP distribution. αSyn-SAA had sensitivity of 97.8% (95% CI 88.4% to 99.9%) in detecting αSyn seeds in neocortical or limbic LRP, but only 14.3% (95% CI 5.0% to 34.6%) in detecting amygdala-predominant LRP (Table 2).

**Table 2.**
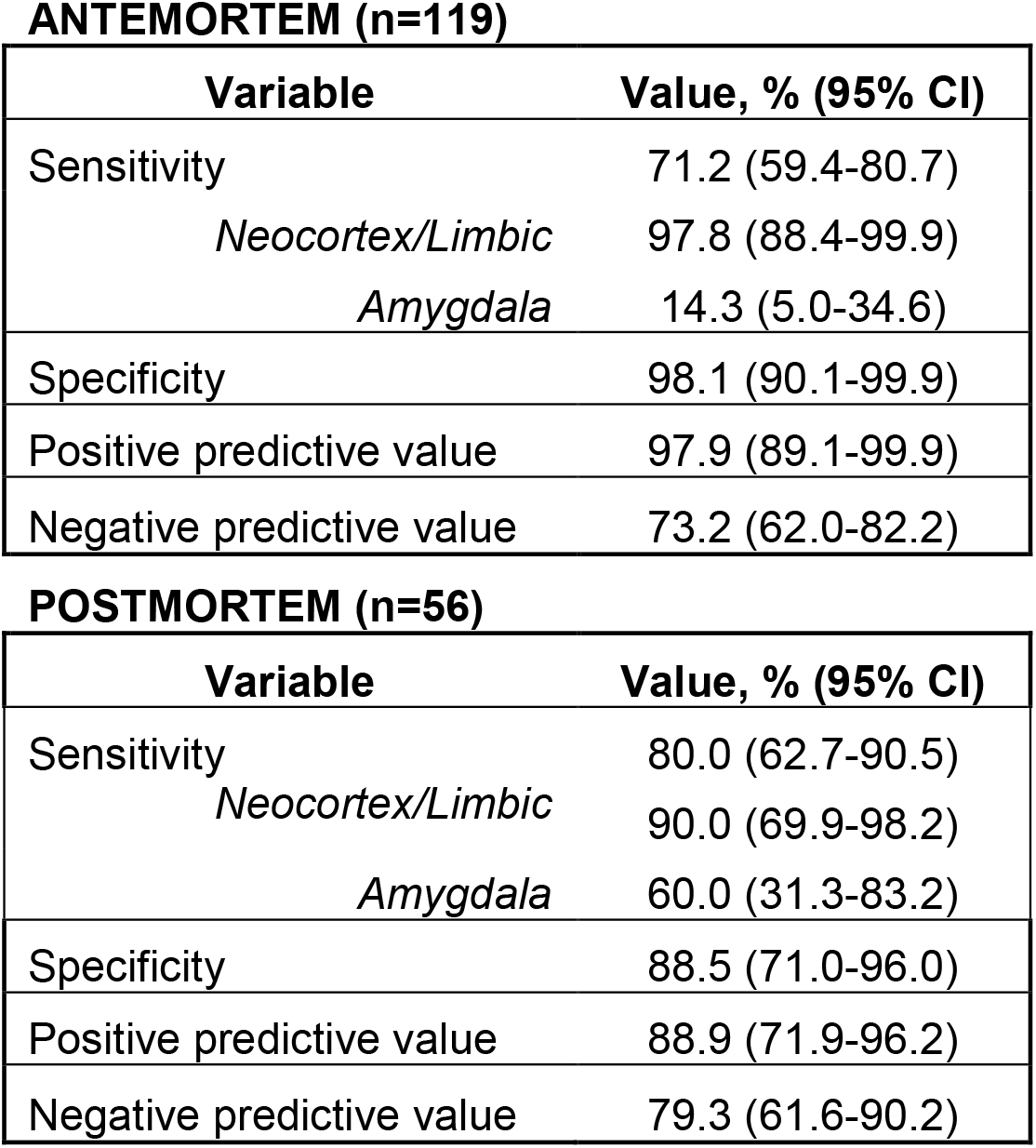
Sensitivity, specificity, and predictive values for antemortem and postmortem CSF αSyn-SAA against LRP.

**ANTEMORTEM (n=119)**
| Variable | Value, % (95% CI) |
| --- | --- |
| Sensitivity | 71.2 (59.4-80.7) |
| *Neocortex/Limbic* | 97.8 (88.4-99.9) |
| *Amygdala* | 14.3 (5.0-34.6) |
| Specificity | 98.1 (90.1-99.9) |
| Positive predictive value | 97.9 (89.1-99.9) |
| Negative predictive value | 73.2 (62.0-82.2) |

**Table 2. Sensitivity, specificity, and predictive values for antemortem and postmortem CSF αSyn-SAA against LRP**
| Variable | Value, % (95% CI) |
| --- | --- |
| Sensitivity | 80.0 (62.7-90.5) |
| *Neocortex/Limbic* | 90.0 (69.9-98.2) |
| *Amygdala* | 60.0 (31.3-83.2) |
| Specificity | 88.5 (71.0-96.0) |
| Positive predictive value | 88.9 (71.9-96.2) |
| Negative predictive value | 79.3 (61.6-90.2) |

Fifty six of the 119 patients had postmortem CSF for αSyn-SAA analysis, 26 who were LRP- and 30 who were LRP+ at autopsy (limbic/neocortical n=20, amygdala-predominant n=10). Of the 26 LRP-patients, 23 were found negative by the clinical αSyn-SAA, for an estimated specificity of 88.5% (95% CI 71.0% to 96.0%) (Table 2). Of the 30 LRP+ individuals, 24 were found positive by αSyn-SAA; thus, the sensitivity for the combined cohort was 80% (95% CI 62.7% to 90.5%). Similarly, when stratified by LRP distribution, the clinical αSyn-SAA in postmortem CSF had sensitivity of 90% (95% CI 69.9% to 98.2%) to detect individuals with limbic or neocortical LRP, but sensitivity of only 60% (95% CI 31.3% to 83.2%), to detect amygdala-predominant LRP (Table 2).

Of the 56 individuals with both antemortem and postmortem CSF, 46 (82.1%) showed concordant αSyn-SAA results, 9 (16.1%) changed from negative results antemortem to positive results on the postmortem assay, and 1 (1.8%) changed from positive to negative. Interestingly, there was a significant difference between LRP groups as to whether or not the αSyn-SAA changed between antemortem and postmortem CSF (*X*^*2*^=28.49, p<0.0001). Of the 10 amygdala-predominant cases with both antemortem and postmortem CSF, 6 changed from false negative to true positive, whereas of the 20 limbic/neocortical cases, only 1 changed αSyn-SAA results. We also found that there was a statistically significant difference in age at death between groups with concordant αSyn-SAA versus αSyn-SAA change (negative to positive), where those with a change had a higher average age at death (F_1,50_=5.44, p=0.02).

116 antemortem (LRP-n=51, limbic/neocortical n=44, amygdala-predominant n=21) and 33 postmortem (LRP-n=11, limbic/neocortical n=15, amygdala-predominant n=7) CSF samples were also analyzed by a kinetic αSyn-SAA to accurately estimate kinetic parameters and further characterize seeding activity in these samples. Fewer samples were run using this assay because some samples had been exhausted in the previous analysis. The kinetic assay provides a diagnostic output based on a probabilistic algorithm, which deems samples as “negative”, “positive”, or “inconclusive. The kinetic αSyn-SAA “negative” and “positive” determinations were consistent with the CLIA-regulated version of the assay for the antemortem and postmortem analyzed in parallel (data not shown). F_max_ was analyzed between LRP groups, with antemortem LRP-(p<0.0001, q=20.42, DF=113) and amygdala-predominant LRP (p<0.0001, q=14.07, DF=113) groups having on average a significantly lower F_max_ than individuals with neocortical or limbic LRP, most likely caused by the abundance of “negative” samples (Figure 2A). Within the antemortem kinetic αSyn-SAA positive patients, the amygdala-predominant group showed a significantly higher time to threshold (TTT) (t(42)=3.681, p=0.0007) and time to reach 50% F_max_ (T_50_) (t(42)=4.033, p=0.0002) compared to the limbic and neocortical group (Figure 2B, 2C). There were no significant differences in kinetic parameters between LRP groups using postmortem CSF in the kinetic αSyn-SAA (data not shown).

**Figure 2.**
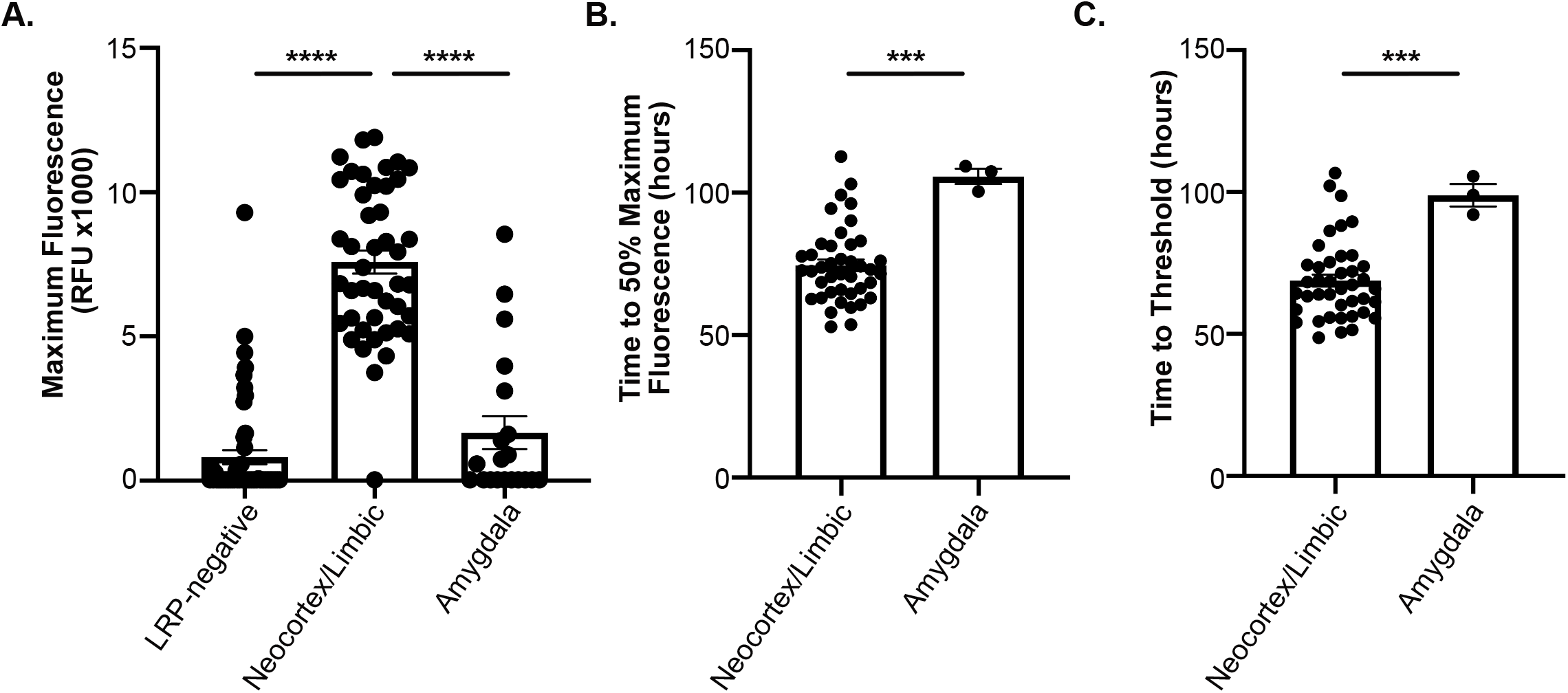
Kinetic parameters of Research SAA stratified by alpha-synuclein distribution. A) Maximum Fluorescence Signal from R/D αSyn-SAA using antemortem CSF between LRP-negative, Neocortex/Limbic, and Amygdala-predominant groups. All samples were included. B) Time to 50% maximum fluorescence and C) Time to Threshold in hours from R/D αSyn-SAA using antemortem CSF between Neocortex/Limbic and Amygdala-predominant groups. “Positive” samples were used. Statistical analysis using one-way ANOVA with Tukey’s multiple comparisons post hoc (A) or unpaired t test (B, C). Error bars represent Standard Error of the Mean (SEM).

### Comparisons of subjects with positive vs negative CSF αSyn-SAA results

We next compared clinical information between the true positive and false negative groups from the antemortem CSF evaluation using the clinical αSyn-SAA. UPDRS part III scores at the time of lumbar puncture were significantly lower in the false negative group compared to the true positive group (Z=-3.12, p=0.002, Fig 3A). The interval between lumbar puncture and death was also significantly different between true positive and false negative groups, with the false negative group having on average a longer interval than the true positive group (Z=2.09, p=0.04, Fig 3B). The two groups also differed in the distribution of Lewy body pathology (*X*^*2*^=48.69, p<0.0001); 94.7% of the false negatives fell into the amygdala-predominant group, while 93.6% of the true positives fell into the limbic/neocortical group (Fig 3C). Similarly, in analyzing postmortem CSF, we found a significant difference across LRP categories in true positive and false negative groups (*X*^*2*^=3.75, p=0.05). Of the false negatives, 66.7% fell into the amygdala-predominant group, while 75% of the true positives fell into the limbic/neocortical group (Fig 3C).

**Figure 3.**
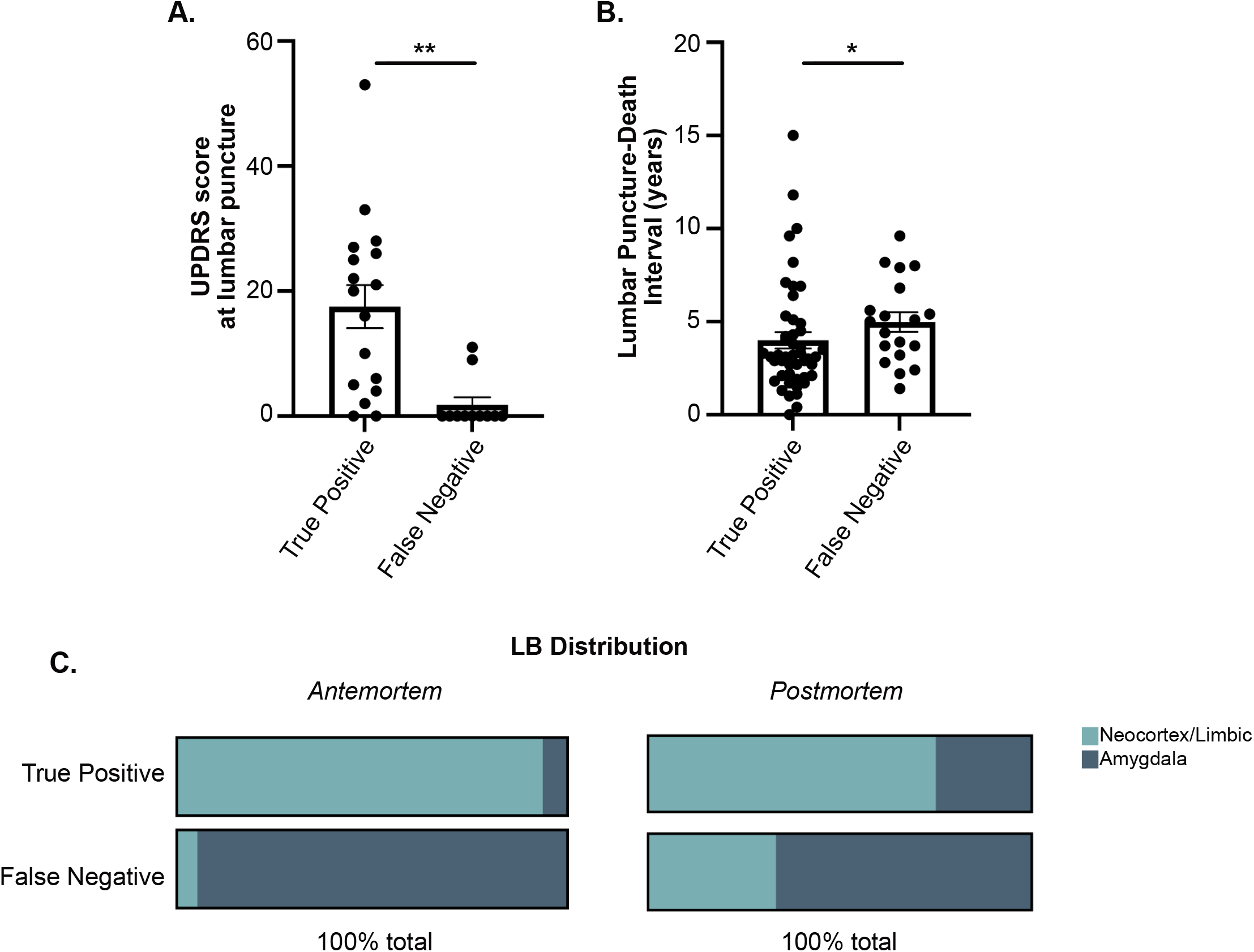
Clinical and pathological differences between True Positive and False Negative. A) UPDRS score at lumbar puncture between True Positive and False Negative groups. B) Interval in years from lumbar puncture to death between True Positives and False Negative groups. C) Distribution of Neocortex/Limbic and Amygdala-predominant LRP in True Positive and False Negative groups for antemortem and postmortem CSF analysis. Statistical analysis using Wilcoxon rank-sum test with post hoc pairwise comparisons from Dwass, Steel, Critchlow-Fligner method (A, B) or chi-square (C). Error bars represent Standard Error of the Mean (SEM).

### Clinical significance of incidental synuclein pathology

Lastly, we explored how clinical diagnosis related to clinical αSyn-SAA performance, in order to better understand whether subtle clinical predictors were present among patients with a diagnosis of a synucleinopathy whose antemortem CSF tested positive by αSyn-SAA. In this analysis, we examined all patients who were clinically diagnosed with AD, without concomitant PF or DLB, and whose αSyn-SAA results were positive versus negative. There was a significant difference in patient biological sex, where αSyn-SAA-positive patients had a significantly greater proportion of males (23/29, 79.3%) compared to αSyn-SAA-negative patients (25/46, 54.3%) (*X*^*2*^=7.84, p=0.005). Clinically diagnosed AD patients with αSyn-SAA-positive CSF had higher UPDRS part III scores than those with αSyn-SAA-negative CSF at most recent visit prior to death (Z=2.53, p=0.01).

### Detection of αSyn seeds from frontal cortex and amygdala brain samples

We next analyzed a subset of patients from the UCSD-ADRC cohort who had frozen brain tissue available for analysis (n=22, LRP-n=4, amygdala-predominant LRP n=10, limbic/neocortical LRP n=8). In both brain regions, the 4 LRP-patients were negative by the αSyn-SAA, consistent with the results for antemortem CSF in both kinetic and clinical assays (Table 3). In agreement with the high sensitivity in CSF for limbic/neocortical LRP cases, seeding activity was detected in both frontal cortex and amygdala of all 8 analyzed cases. However, there was a significant decrease in seeding activity in both frontal cortex and amygdala of the amygdala-predominant LRP cases. Of the 10 amygdala-predominant cases, 4 cases showed no seeding activity in both frontal cortex and amygdala. There were 2 cases with seeding activity detected in the amygdala, with one of them showing 2/3 wells positive in frontal cortex.

**Table 3.**
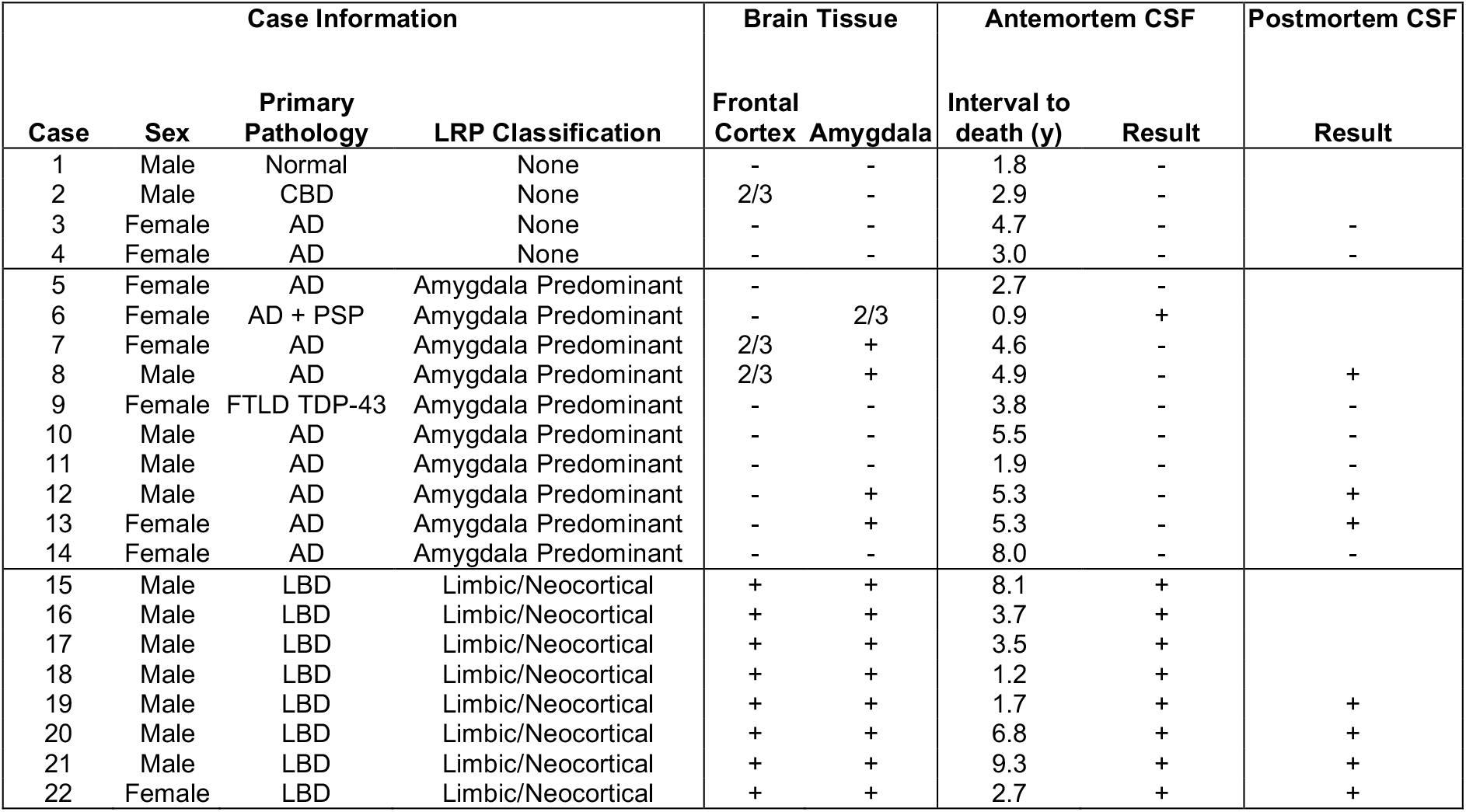
Patient categorization from brain homogenate samples. Abbreviations: NA: not applicable. LRP: Lewy Related Pathology, CSF: cerebrospinal fluid. CBD: corticobasal degeneration, AD: Alzheimer’s disease, PSP: progressive supranuclear Palsy, FTLD TDP-43: frontotemporal lobar degeneration TAR DNA-binding protein 43, LBD: Lewy-body disease. Inconclusive cases have 2/3 replicate wells that were positive. Brain tissue samples were analyzed at 10^−8^ dilution. Positive results indicate 3/3 replicates were positive and negative results indicate 0 or 1 out of 3 replicates were positive. Amygdala tissue could not be obtained for case 5.

Within LRP- and limbic/neocortical LRP groups, there was 100% concordance between brain homogenate results and CSF results. Of the 5 patients with amygdala-predominant LRP that also tested positive on the αSyn-SAA using amygdala brain tissue, 3 also had some seeding activity on the αSyn-SAAs using either antemortem or postmortem CSF (Table 3). Overall, the assay detected higher seeding activity in amygdala tissue in amygdala-predominant cases, while neocortical cases presented high levels of seeding activity in both brain regions.

## Discussion

Although there have been large strides in the understanding of the molecular basis of synucleinopathies, *in vivo* methods for detecting αSyn are still limited. Misfolded αSyn aggregation likely begins years to decades before the onset of symptoms, allowing for the potential ability to identify patients in the earliest stages of their diseases. The development of a sensitive and specific diagnostic tool for synucleinopathies would allow for early diagnosis of patients where often there is the highest level of clinical uncertainty and when disease modifying therapies are of the greatest potential use^35^. Furthermore, the ability to detect both primary and secondary αSyn pathologies may have ramifications not only for prognostication, but also for the enrollment into protein-directed clinical trials. Thus, αSyn-related biomarkers remain a crucial need to the field. Several publications have shown promising results for αSyn-SAAs (αSyn-PMCA and RT-QuIC) performed in academic laboratories^16, 36^, but the performance of the assay within a regulated CLIA environment, and against pathology-confirmed samples, has not been evaluated until now. Most studies have focused on evaluations of accuracy for diagnostic purposes, although there are indications that αSyn-SAA results correlate with pathological and/or clinical findings. F_max_ correlates with αSyn seed structure, effectively differentiating PD from MSA^24^, and faster amplification has been observed for DLB compared to PD^37^. Thus, αSyn-SAA evaluation of samples from patients with mixed clinical diagnoses and mixed pathological findings may offer new insights in antemortem diagnosis and disease management. Moreover, the knowledge of whether current generations of αSyn-SAAs can detect these secondary pathologies is crucial to understanding the range of their diagnostic utility. Therefore, we studied a large autopsy-validated multicenter cohort of patients with a spectrum of LRP, including the amygdala-predominant pattern commonly observed in association with AD pathology.

Our results add to the previous reports that αSyn-SAAs can robustly detect αSyn seeds in limbic/neocortical stage LRP, but also show decreased sensitivity in detecting αSyn seeds in amygdala-predominant disease. An additional unique feature to this study is the number of subjects with postmortem CSF, providing a proximal time point to the autopsy assessment. Classification using postmortem CSF showed a sensitivity of 80% and specificity of 88.5%, however when stratified by pathology distribution, again the assay performed significantly better in detecting limbic/neocortical LRP than amygdala-predominant LRP. Lastly, we also observed decreased seeding activity from amygdala-predominant cases when assaying frozen brain tissue from frontal cortex and amygdala.

The lack of concordance between CSF αSyn-SAA and amygdala-predominant pathology may represent assay dependence on the amount of brain LRP “burden”. In agreement with this interpretation, patients with higher levels of seeds in amygdala homogenate were positive in CSF. Moreover, there was no difference in the F_max_ between positive CSF samples from amygdala-predominant and limbic-neocortical cases, like those reported between PD and MSA. Alternatively, negative αS-SAA CSF samples in the amygdala-predominant group could be explained by localized brain pathology that does not enter the CSF. However, direct analysis of amygdala homogenate from amygdala-predominant cases showed low detection, suggesting less seeding activity by these particular αSyn species. Recent studies have found that αSyn species in amygdala-predominant pathology found in AD may have different properties than PD or DLB patients with limbic and neocortical LRP^14, 38, 39^. It is plausible that these amygdala-predominant αSyn seeds have lower rates of amplification than diffuse LRP due to unique conformation of these seeds.

Since αSyn pathology commonly coexists in AD, and may be associated with faster clinical progression^40^, identifying this pathology with a biomarker would improve clinical monitoring and create options for clinical trials targeting αSyn in these patients. If amygdala-predominant type αSyn pathology is an early stage or precursor of more widespread concomitant LB pathology in AD, then detecting its presence through biomarkers such as αSyn-SAA would be useful. However, the effect of amygdala-predominant LRP in AD appears to have less clinical impact in some cases or may take years to convert to a more widespread seeding. Further work is needed to determine why the seeding potential of amygdala-predominant αSyn pathology is lower in some cases, or whether different types of αSyn-SAAs could provide detection of this pathology.

The assay’s ability to identify clinically unexpected synuclein pathology is an area of great potential. Our results indicate that 27/75 (36%) of the clinically diagnosed AD patients had αSyn aggregates in their antemortem CSF and were later autopsy-confirmed to have limbic/neocortical LB disease. DLB is frequently misdiagnosed as AD during life, and the presence of moderate to severe AD-related tau pathology is associated with a lower likelihood of visual hallucinations and cognitive fluctuations, and worse performance on tests of episodic memory and naming in DLB patients, meaning that it is more challenging to diagnose these patients with mixed pathologies accurately^27, 41, 42^. UPDRS III scores were higher amongst those who would test positive and thus detailed motor exams in dementia patients remain essential. Biological sex was also different between these groups, which is somewhat expected as there is an already established relationship between synucleinopathies and the male sex^43, 44^. These findings may become useful when establishing a criterion for when to undergo lumbar puncture to confirm the presence of a synucleinopathy in dementia patients in the clinical setting.

Some limitations to this study include modest differences in the clinical and pathological assessments between the two institutions, although each adhered to NACC guidelines. The difference in time from CSF collection to autopsy between the true positive and false negative cases, suggests that the false negative group is most likely inflated by cases in which LRP became detectable after antemortem CSF sampling. This is consistent with the older age at death for the individuals that changed from false negative to true positive between antemortem and postmortem CSF analysis. Additionally, there were no brainstem-only LB cases in our patient cohort. Future studies should include assessments in larger population-based cohorts and use different αSyn-SAAs to determine if this finding is generalizable beyond the current studied setting. Lastly, further work is needed to more fully interrogate differences in αSyn species between these groups that may account for the variability in seeding capability.

In this large, multicentered autopsy-validated cohort of patients with a variety of stages of LRP, our results indicate that the αSyn-SAA is highly predictive of neocortical or limbic LRP in aging patients for whom Lewy bodies are not clinically suspected, making it a diagnostic tool with great potential for clinical trials aiming to initiate interventions early in the disease process. However, there was substantially lower sensitivity to detect amygdala-predominant LRP in brain tissue and CSF, which is often observed in the setting of AD pathology and may have distinct biochemical properties and seeding potential that affect the current generation of αSyn-SAAs.

## Data Availability

All data produced in the present study are available upon reasonable request to the authors.

## Acknowledgments

We thank the patients who participated in this research, their families, and the investigators and staff at the OHSU Layton Aging and Alzheimer’s Disease Research Center and Oregon VA Parkinson’s Disease Research Education and Clinical Care Center; UCSD Shiley Marcos Alzheimer’s Disease Research Center; Robin Guariglia and Nora Mattek for compilation of clinical and genetic data; Babett Lind for CSF sample collection. Additional thanks to the individuals at Amprion Inc. involved in this project: Frank Espin, John Middleton, Russell M. Lebovitz, and Karen MacLeod.

This study was funded in part by the Alzheimer Disease Center Clinical Core at Oregon Health and Science University (PI: Kaye; eIRB 725; supported by NIH P30 AG008017, P30 AG066518) as well as the National Center for Advancing Translational Sciences (National Institutes of Health, Grant Award Number UL1TR002369). Amprion’s efforts were funded in part by the Alzheimer’s Drug Discovery Foundation (ADDF) Diagnostics Accelerator, as well as by the National Institute of Neurological Disorders and Stroke of the National Institutes of Health (Award Number U44NS111672). Moriah R. Arnold is funded through the Medical Scientist Training Program of Oregon Health & Science University (T32 GM 109835). Dr. Coughlin is funded by the National Institute of Neurological Disorders and Stroke of the National Institutes of Health (Award Number NS120038) and the National Institute on Aging (Award Number AG062429). Dr. Galasko is funded by the UCSD Shiley-Marcos ADRC (AG062429), and by the DLB Research Center of Excellence award from the Lewy Body Dementia Association. The funding sources had no role in the design and conduct of the study; collection, management, analysis, and interpretation of the data; preparation, review of approval of the manuscript; and decision to submit the manuscript for publication.

## Author Contributions

MRA and DGC designed studies, analyzed data, and participated in the writing of the manuscript. BHB ran the statistical analysis of data. DSS participated in designing the studies and editing the manuscript. LCM, CMF, and YM participated in the research and development assay used for brain homogenates and CSF. AH, DCG, and YK were responsible for neuropathology analysis at UCSD. JAK was involved in the collection of patient samples at the OADC. RLW was responsible for neuropathology analysis at OHSU. LCM, DRG and JFQ designed studies, helped interpret data, and participated in the writing of the manuscript. All authors participated in critical review of the manuscript. MRA, BHB, and JFQ had full access to all the data in the study and takes responsibility for the integrity of the data and the accuracy of the data analysis.

## Potential Conflicts of Interest

Dr. Concha, Ms. Farris, and Mr. Ma are inventors on several patents related to PMCA technology (SAA) and are associated to Amprion Inc, a biotech company focused on the commercial utilization of SAA for diagnosis. All other authors have no conflicts of interest to disclose.

## Notes

### Author Declarations

eIRB 725 of Oregon Health and Science University ADRC gave ethical approval for this work. IRB 170957 of University of California San Diego ADRC gave ethical approval for this work.

